# Association of trajectories of depressive symptoms with vascular risk, cognitive function and adverse brain outcomes: The Whitehall II MRI sub-study

**DOI:** 10.1101/2020.05.20.20106963

**Authors:** N Demnitz, M Anatürk, CL Allan, N Filippini, L Griffanti, CE Mackay, A Mahmood, CE Sexton, S Suri, AG Topiwala, E Zsoldos, M Kivimäki, A Singh-Manoux, KP Ebmeier

**Affiliations:** Department of Psychiatry, University of Oxford, Oxford, UK; Global Brain Health Institute, Trinity College Dublin, Dublin, Ireland; Institute of Translational and Clinical Research, Newcastle University, UK; Cumbria, Northumberland, Tyne and Wear NHS Foundation Trust, UK; Oxford Centre for Human Brain Activity, Wellcome Centre for Integrative Neuroimaging, Department of Psychiatry, University of Oxford, UK; Department of Population Health, London School of Hygiene and Tropical Medicine, London, UK; Department of Epidemiology and Public Health, University College London, London, UK; Université de Paris, INSERM U1153, Paris, France

**Keywords:** Depressive symptoms, aging, white matter hyperintensities, cognition, late onset, vascular

## Abstract

**Background:** Trajectories of depressive symptoms over the lifespan vary between people, but it is unclear whether these differences exhibit distinct characteristics in brain structure and function.

**Methods:** In order to compare indices of white matter microstructure and cognitive characteristics of groups with different trajectories of depressive symptoms, we examined 774 participants of the Whitehall II Imaging Sub-study, who had completed the depressive subscale of the General Health Questionnaire up to nine times over 25 years. Twenty-seven years after the first examination, participants underwent magnetic resonance imaging to characterize white matter hyperintensities (WMH) and microstructure and completed neuropsychological tests to assess cognition. Twenty-nine years after the first examination, participants completed a further cognitive screening test.

**Results:** Using K-means cluster modelling, we identified five trajectory groups of depressive symptoms: consistently low scorers (“low”; n=505, 62.5%), a subgroup with an early peak in depression scores (“early”; n=123, 15.9%), intermediate scorers (“middle”; n=89, 11.5%), a late symptom subgroup with an increase in symptoms towards the end of the follow-up period (“late”; n=29, 3.7%), and consistently high scorers (“high”; n=28, 3.6%). The late, but not the consistently high scorers, showed higher mean diffusivity, larger volumes of WMH and impaired executive function. In addition, the late subgroup had higher Framingham Stroke Risk scores throughout the follow-up period, indicating a higher load of vascular risk factors.

**Conclusions:** Our findings suggest that tracking depressive symptoms in the community over time may be a useful tool to identify phenotypes that show different etiologies and cognitive and brain outcomes.

---

Depressive symptoms, when tracked longitudinally in the general population, show heterogeneous patterns of change over time. Although most individuals experience few or no symptoms, some experience them transiently, while others consistently experience depressive symptoms (1). These different trajectory patterns are hypothesized to have different implications for brain structure and function in late adulthood, but evidence for this hypothesis is scarce.

A number of studies have used trajectory analyses to identify groups based on longitudinal patterns of change in depressive symptoms (for review, see (1)). For example, Mirza et al. (2) tracked depressive symptoms over the course of 11 years in a sample of 3,325 older adults and identified 5 trajectories: “low”, “increasing”, “decreasing”, “remitting” and “high”. The study showed that the trajectory characterized by increasing depressive symptoms was associated with a higher risk of developing dementia (2). Similarly, in an analysis of depressive trajectories over 28 years, a late-onset, but not early-onset of depressive symptoms was predictive of an increased dementia risk (3). Almeida et al. (4) described a greater risk of dementia over 14 years follow-up in 4922 cognitively healthy 71 to 89-year-olds with current depression, as opposed to those with a past history of depression or no history of depression. These findings have led to the suggestion that depressive symptoms are a prodromal feature of dementia, or that the two share common causes, as opposed to depression being a causal risk factor for dementia (2, 3, 5). However, it remains unclear whether the groups that emerge directly from trajectory modelling based on depressive symptoms can also be predictive of cognitive impairment and structural brain differences.

The vascular depression hypothesis proposes that comorbid late onset depression (LOD) and cerebrovascular disease disrupts fiber tracts within prefrontal systems (6). A meta-analysis of studies investigating white matter changes in late life depression found that LOD was characterized by more frequent and intense white matter abnormalities (5). Longitudinal studies are required to test the relationship between depressive symptom trajectories and structural abnormalities directly, as well as their role in cognitive function and deterioration.

We hypothesize that temporal patterns of depressive symptoms have implications for cognitive function. While greater cognitive impairment has been reported in patients with LOD compared to those with early onset depression (EOD) (7, 8), the reverse pattern (9), as well as no group differences (10), have also been noted. In addition, the group-difference between LOD and EOD may be specific to cognitive domains. Interestingly, a review of studies comparing cognitive outcomes between EOD and LOD groups found that patients with LOD showed greater reductions in executive function than patients with EOD and controls, while memory impairments were equivalent in the two groups (7).

We hypothesize that participants showing a trajectory of late-onset depressive symptoms have a larger volume of white matter hyperintensities and indices of impairments in white matter microstructure, such as greater mean diffusivity and reduced fractional anisotropy. In line with our previous findings (7), we expect the late-onset group to have reduced executive function and be at greater risk of further cognitive deterioration.

## Methods

### Study sample

At baseline, the Whitehall II Study recruited, and continues to follow-up, a cohort of 10,308 participants who worked in the British civil service in 1985 (11). Eight hundred of these were randomly selected from Wave 11 (2012/13, see Figure 1) and recruited for the Whitehall II Imaging sub-study (12). We included participants who completed at least four of nine instances of the General Health Questionnaire (GHQ) depressive subscale during the follow-up period (Figure 1), as well as a 3T MRI brain scan and a battery of cognitive tests on average 27.3 (SD = 1.5) years after the first GHQ assessment. We excluded from analyses participants who reported a history of neurological illness (participants who reported a history of transient ischemic attacks were included) or displayed significant abnormalities on structural MRI scans. Participants who had no available DTI or WMH data were excluded only from the analyses of that outcome. Thus, the total analytic sample from which the trajectories were derived included 774 participants, while analyses of WMH and DTI outcomes had 750 and 728 participants, respectively.

**Figure 1.**
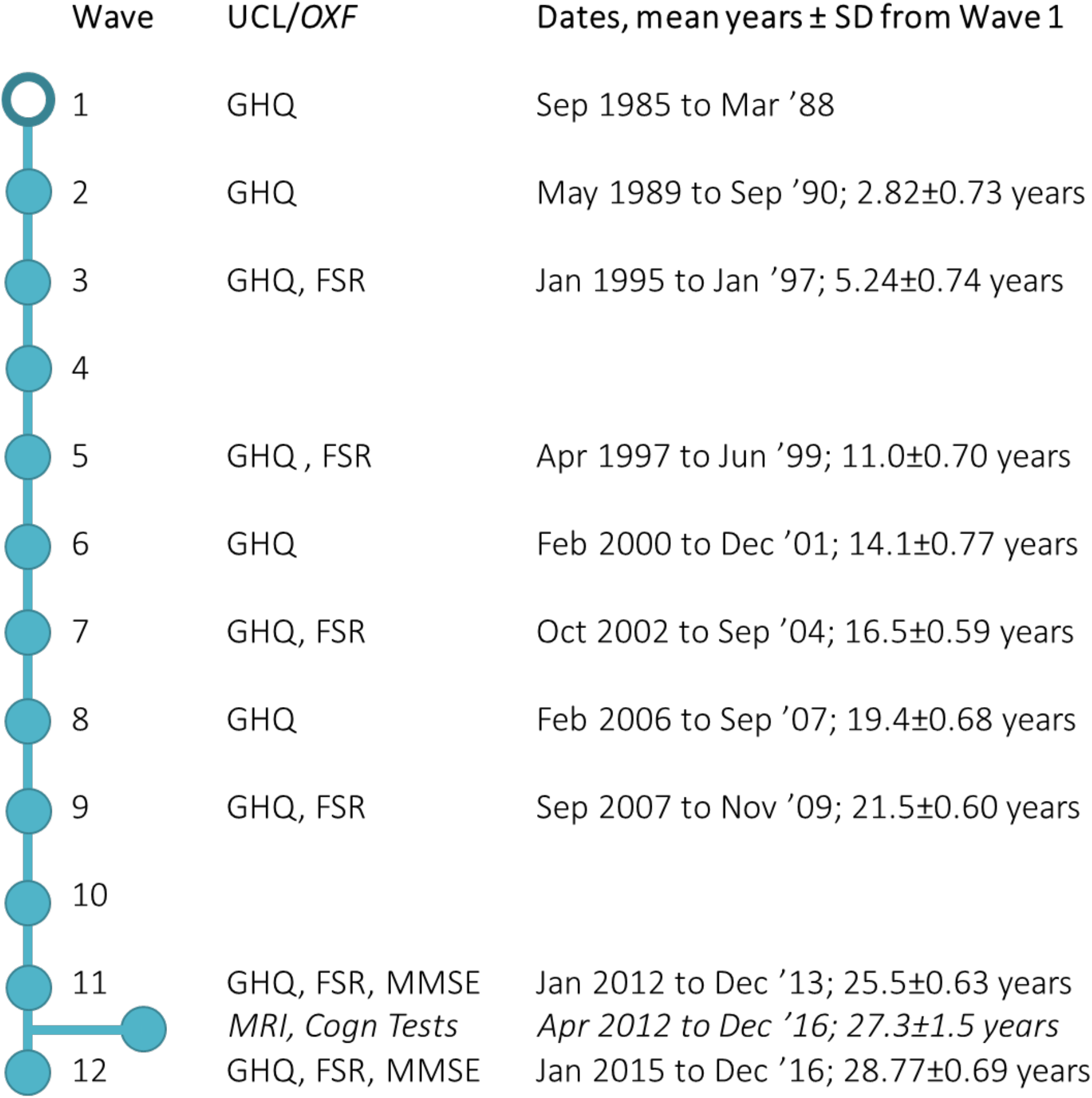
Timeline of assessments. *GHQ (General Health Questionnaire), MMSE (Mini Mental State Examination), FRS (Framingham Stroke Risk score), BP (mean arterial blood pressure), MRI (Magnetic Resonance Imaging), COG (Cognitive Test battery described in (12)). All measurements were acquired at University College London, except for those indicated in italics, which were obtained at the University of Oxford*.

The Whitehall II Study and the Whitehall II Imaging sub-study received ethical approval from the University College London Medical School Committee on the Ethics of Human Research (Reference: 85/093) and the Oxford Central University Research Ethics Committee (Reference: MS IDREC-C1-2011-71), respectively. Informed written consent was obtained from all participants at all waves.

### Depressive symptoms

Depressive symptoms were measured as total score, ranging from 0 to 12, on the depressive subscale of the General Health Questionnaire (GHQ). The GHQ has previously been validated for Whitehall II data and shown to have good criterion validity for minor psychiatric disorders (13). Participants completed the GHQ at nine waves from 1985-2013 (Figure 1).

### Neuropsychological Tests

At the time of the MRI scan, participants completed a battery of cognitive tests (12). In line with our hypothesis, and to limit multiple comparisons, only tests of memory and executive function were included here in addition to a global cognitive screening test. For the measurement of executive function we selected the Trail-Making Test (Completion Time Part B – Part A) (14). Memory performance was measured using the total learning score over three trials of the Hopkins Verbal Learning Test Revised (HVLT-R) (15). The Mini Mental State Examination (MMSE) was used to assess global cognitive function in Waves 11 and 12 (16).

### Vascular risk

Vascular risk was assessed using the Framingham stroke risk scores (in percent risk over the following 10 years) collected during Waves 3, 5, 7, 9, and 11 and log10-transformed for analysis. Mean arterial BP at the time of scan was computed as the weighted mean (1:2) of systolic and diastolic BP.

### MRI acquisition and processing

As previously described (12), magnetic resonance imaging data were acquired on a 3T Siemens Magnetom Verio scanner in the first 550 participants (between April 2012 – December 2014), using a 32-channel head coil, at the Oxford Centre for Function MRI of the Brain. T1-weighted structural images were acquired using a gradient echo sequence (TR = 2530ms, TE= 1.79/3.65/5.51/7.37ms, flip angle = 7°, FOV = 256mm, voxel dimension = 1.0 mm isotropic, acquisition time = 6m12s). Diffusion-weighted images were acquired using an echoplanar sequence, with 60 diffusion-weighted directions (*b*-value = 1500 s/mm2), 5 non-diffusion weighted images (*b*-value = 0s/mm2) and one b0 volume in the reversed phase-encoded direction (TR = 8900ms, TE = 91.2ms, FOV= 192 mm, voxel dimension = 2.0mm isotropic). FLAIR images were also acquired (TR = 9000ms, TE = 73, voxel dimension=0.9×0.9×3mm^3^, FOV = 220, acquisition time = 4m14s). The last 250 participants were scanned on a 3T Siemens Magnetom Prisma scanner with 64-channel receive head-neck coil in the same center (between July 2015 – December 2016). T1-weighted structural images were acquired using a gradient echo sequence (TR = 1900ms, TE= 3.97ms, flip angle = 8°, FOV = 192mm, voxel dimension = 1.0 mm isotropic, acquisition time = 5m31s). FLAIR (TR = 9000ms, TE = 73, voxel dimension=0.4×0.4×3mm^3^, FOV = 220, acquisition time = 4m14s) and diffusion-weighted images, with a matched protocol except for a change in echo time (TE = 91ms), were also acquired. Analysis of MRI data was conducted using tools from the FMRIB Software Library (FSL). Partial-volume tissue segmentation was performed using the FMRIB Automated Segmentation Tool (FAST)(17). Diffusion-weighted images were first corrected for head movement and eddy currents. Global measures of fractional anisotropy (FA) and mean (MD) diffusivity were then extracted using DTIFit (http://fsl.fmrib.ox.ac.uk/fsl/fdt). First, FA maps were created by fitting a tensor model to the raw diffusion data. This fits a diffusion tensor model to the raw diffusion data and then applies BET to remove non-brain voxels (18). All participants’ FA data were aligned into standard space using FMRIB’s Nonlinear Registration Tool (FNIRT) (19). The mean FA image was then calculated and thinned to create a mean FA skeleton representing the centers of all tracts common to the group. This method was repeated for mean diffusivity (MD). Measures of FA and MD were then extracted from the TBSS skeleton.

To acquire an estimate of white matter hyperintensity volume, FLAIR images were automatically segmented using the Brain Intensity AbNormality Classification Algorithm (BIANCA), a fully-automated, supervised method for WMH detection, based on the k-nearest neighbor algorithm (20). Briefly, BIANCA classifies the image voxels based on their intensity and spatial features. The output image represents the probability per voxel of being WMH. In order to adjust for brain volume, total WMH volume was then divided by total intracranial volume (ICV) and multiplied by 100 (WMH%). The application of this processing pipeline to this dataset has been described elsewhere (21). All analyses of imaging data included a binary covariate for scanner.

### Sample characteristics and covariates

Age, sex and years of full-time education were recorded for all participants at the time of the MRI. Social-economic stratum was determined according to occupation in 1985–88 (Wave 1). At the time of the scan (2012-16), premorbid full-scale intelligence quotient (FSIQ) was estimated from the Test of Premorbid Function (TOPF), and the Pittsburgh Sleep Quality Inventory (PSQI) was used to measure sleep quality (22). To describe the diagnostic validity of the depressive symptom trajectories, the following measures, also collected at about the time of scan, were used: ICD10 History of depressive episode determined from CIS-R in 2012-13 (13), DSM IV-TR criteria for lifetime history of minor and major depressive episodes diagnosed by SCID in 2012-16 (23), self-reported clinical diagnosis of depression and current treatment with antidepressant drugs at the same time, as well as the Centre for Epidemiological Studies Depression Scale (CES-D) (24).

### Statistical analysis

K-means cluster modelling (kml) was used to identify distinct cluster trajectories of GHQ depression scores. Kml is a non-parametric hill-climbing algorithm which does not impose assumptions regarding the shape of the trajectories (25). The kml analysis was specified to allow between 2 and 5 clusters (trajectories), each obtained by running 1000 permutations. The Calinski and Harabasz criteria, along with consideration of clinical relevance, were used to select the optimum number of clusters. Kml offers 3 versions of the Calinski and Harabasz criteria, described in detail elsewhere (26), and all were considered. While the traditional and Kryszczuk variants of the Calinski and Harabasz criterion indicated that the optimum number of clusters was 2, the Genolini variant suggested a 5-group result. The 2-cluster response grouped participants into those who consistently reported minimal depressive symptoms (81%), and those who showed some symptoms (19%). Although this clustering resulted in a higher Calinski and Harabasz criterion in 2 variants, we opted for the 5-cluster response, because (a) the 2-groups solution is of limited clinical relevance as it combines all trajectories presenting some depressive symptoms, thus losing important information about their trajectories of change, and (b) our research question addresses a group showing a later emergence of depressive symptoms. K-means cluster modelling for longitudinal data and statistical analyses were performed in R version 3.5.1 with RStudio version 1.1.463 (27) using the psych (28), kml (25) and ggplot2 packages (29).

Data were inspected by plotting variables by trajectory group, in particular for variables that replicate and validate the depression measures, i.e. CES-D, as well as symptoms scales related to clinical depression, such as the Pittsburgh Sleep Quality Inventory, and physiological measures, such as mean arterial blood pressure. Similarly, cognitive variables that we predicted to be abnormal in a late-onset trajectory were inspected in the same way: MMSE, HVLT-R, and the difference measure between Trails A and B (TMT; representing executive function). Finally, following the vascular hypothesis of late onset depression, we plotted group means of Framingham Stroke risk scores computed in Waves 3, 5, 7, 9, and 11. All measures were adjusted for age and sex differences between trajectory groups. In addition, group differences in cognitive outcomes were also adjusted for premorbid IQ.

### Adverse cognitive and brain outcomes

Hypothesis testing was carried out for brain measures at the MRI phase, 27.3 (SD = 1.5) years after the first GHQ assessment. ANCOVAs were then used to evaluate whether the identified trajectory groups significantly differed on cognitive and brain structure outcomes. In addition, we tested whether group membership was associated with follow-up MMSE scores, measured 28.8 years (SD: 0.69) after the first examination. Age, sex, education and premorbid IQ were included as covariates in the analysis of cognitive data. Age, sex, education, premorbid IQ, and MRI scanner were included as covariates in the analysis of MRI data. Planned post-hoc tests between the late onset and all other trajectory groups were carried out following our primary hypothesis.

## Results

A total of 774 participants were included in the analyses. Our sample was mostly male (81%), had a mean age of 69.8 ± 5.2 years and had, on average, pursued 14.8 ± 3.3 years of full-time education.

The K-means cluster analysis identified five distinct trajectories of depressive symptoms in these 774 participants (Figure 2a). The majority of participants maintained a low score of depressive symptoms throughout the study (low depressive symptoms; n = 505, 65.2%). The remaining four trajectories were characterized by early depressive symptoms that then decreased (“early” depressive symptoms; n = 123, 15.9%); moderate levels of depressive symptoms gradually decreasing after Wave 7 (“middle” depressive symptoms; n = 89, 11.5%); low starting scores that steadily increased throughout follow-up (“late” depressive symptoms; n = 29, 3.7%); or maintained high scores throughout (consistent “high” depressive symptoms n = 28, 3.6%).

**Figure 2a.**
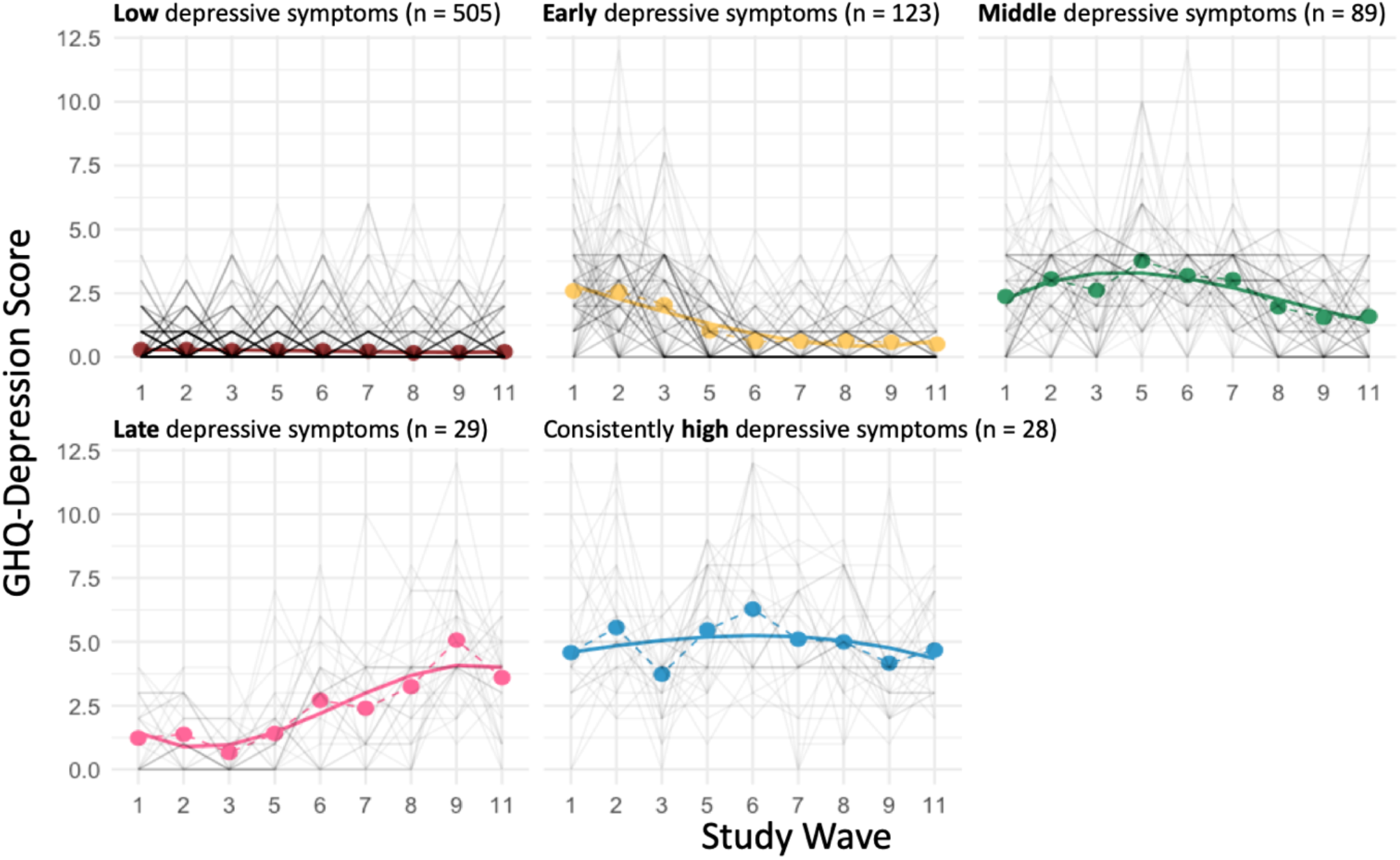
Trajectories of depressive symptoms from Wave 1 (1985-1987) to Wave 11 (2012-2013), with a mean follow-up time of 25.6±0.6 years. The figure shows the five estimated trajectories of depressive symptoms measured in 774 participants using the GHQ-depression subscale. Solid lines indicate the mean trajectory, while the dotted lines connect mean individual scores for each wave.

### Diagnostic validation

Figure 2b shows the percentage of participants qualifying for GHQ-caseness (GHQ depression sub-score>3) in each group by waves. Table 1 describes the trajectory groups in terms of ICD10 History of a depressive episode determined from CIS-R in 2012-13 (13), as DSM IV-TR criteria for lifetime history of minor and major depressive episodes diagnosed by SCID in 2012-16 (23), and self-reported clinical diagnosis of depression and current treatment with antidepressant drugs at the same time.

**Figure 2b.**
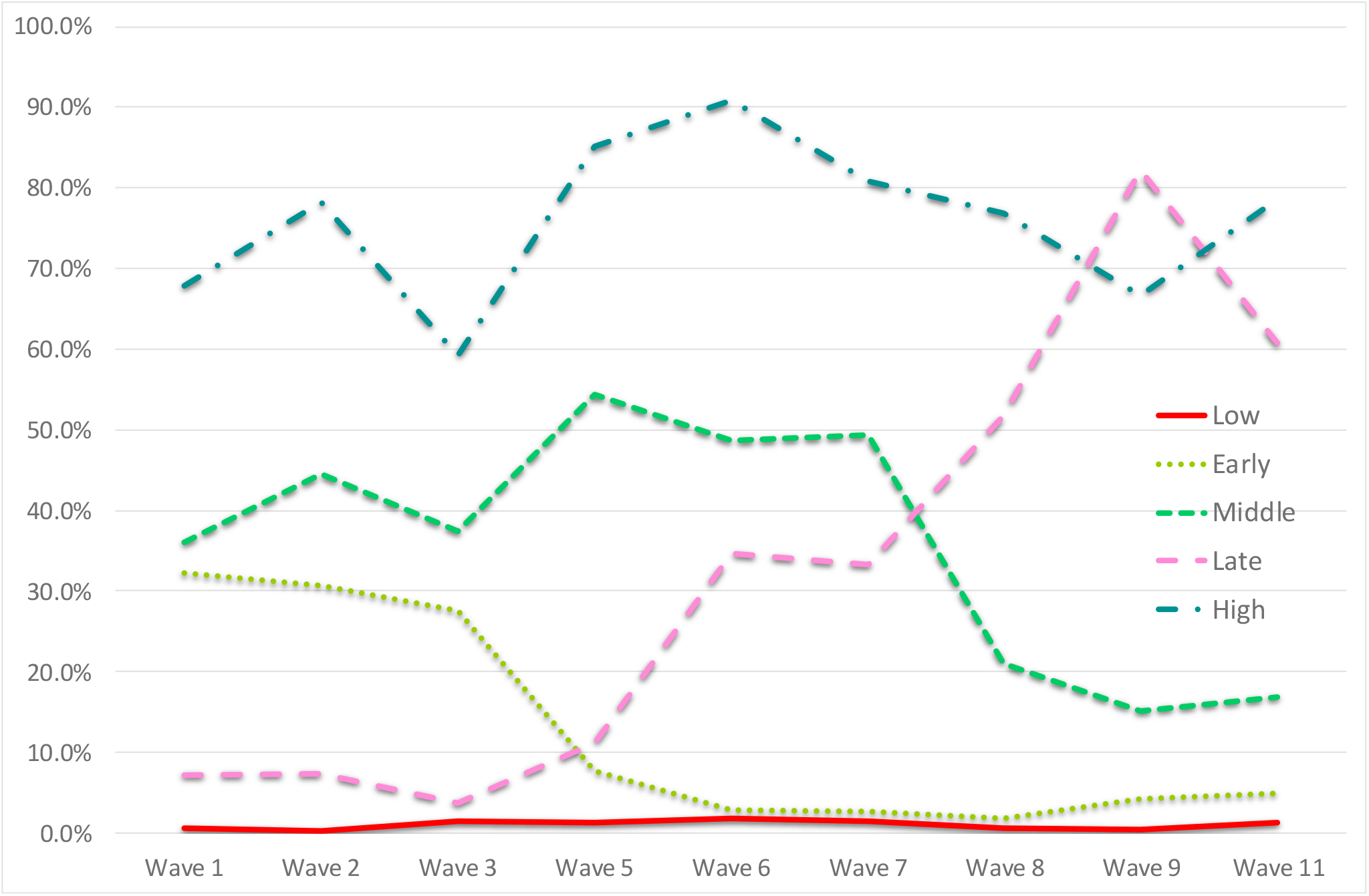
Percent GHQ cases by wave in each depression trajectory group. Cases ≈ GHQ > 3.

**Table 1.**
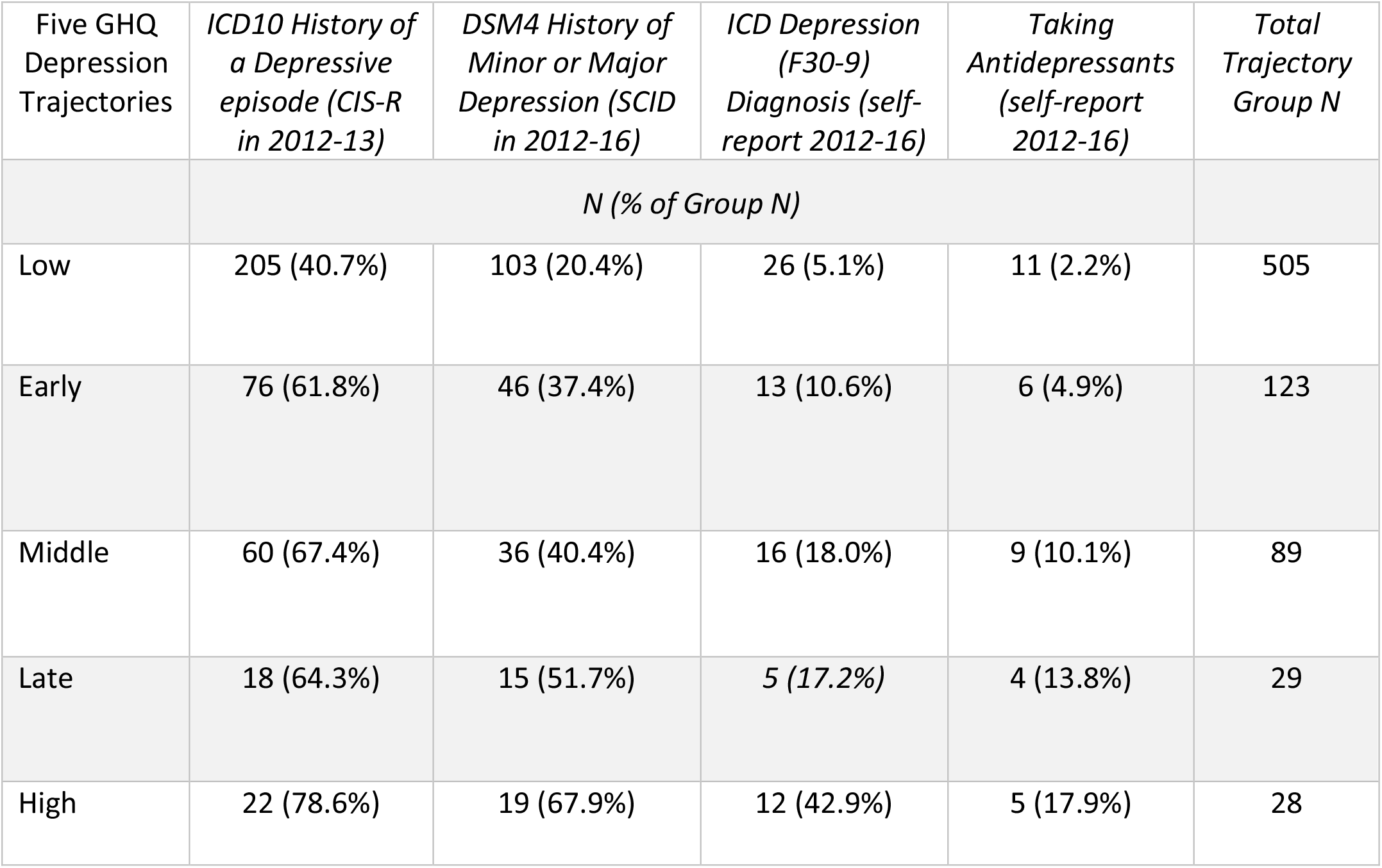
Diagnostic Validation.

### Sample characteristics of trajectory groups

Participants in the five trajectory-based groups did not significantly differ in terms of age at scan, ethnic group (white/other), socio-economic stratum, years of education, or pre-morbid IQ. However, as expected, the “late depressive symptoms” and “consistently high depressive symptoms” groups had a significantly larger proportion of women to men and higher CES-D scores at time of scan compared with individuals in the other trajectories (Table 2).

**Table 2.**
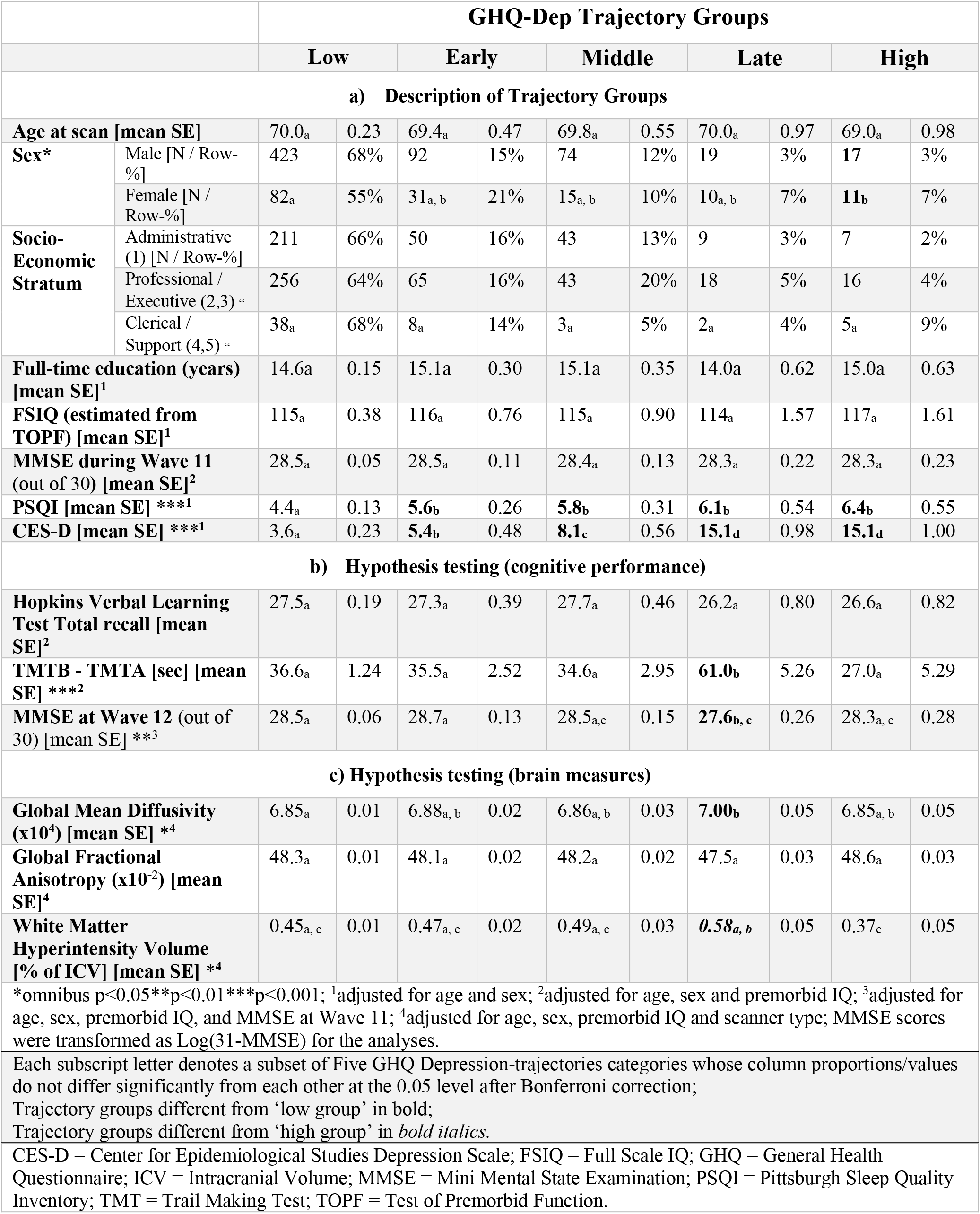
Characteristics of estimated GHQ trajectory groups.

Reflecting the presence of depressive symptoms at the end of the follow-up period, both “late” onset and consistently “high” symptom groups had higher CES-D depression scores than the three other groups, and together with the “early” and “middle” symptom groups they reported poorer sleep quality measured by the Pittsburgh Sleep Quality Inventory than the “low” symptom group. As predicted, and consistent with the vascular depression hypothesis (6), the “late” depressive symptoms group had higher vascular risk (Framingham Stroke Risk Score) on average during the preceding 28 years, and individually (Bonferroni-controlled) as far back as 22 years during Wave 3, as well as a higher mean arterial BP at the time of the scan (Table 3).

**Table 3.**
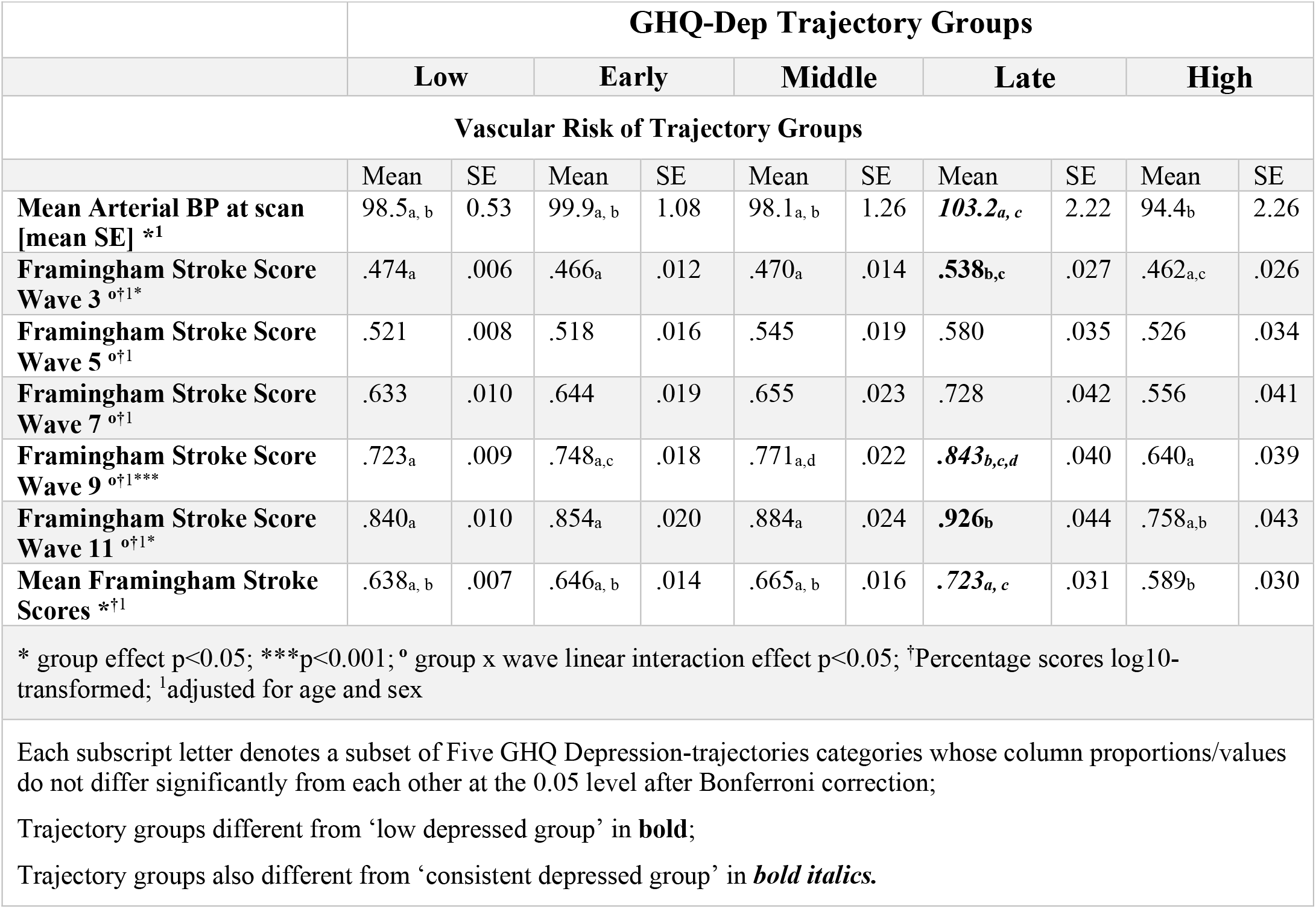
Vascular risks in estimated GHQ trajectory groups.

### Adverse cognitive and brain outcomes

ANCOVAs indicated that the “late” depressive symptom group had a poorer performance on the executive component of the Trail Making Test, higher mean diffusivity, and a significant deterioration of MMSE score between Waves 11 and 12 (Table 2, Figure 4). Performance on the HVLT-R (total recall) and MMSE at the time of scan did not differ between groups. While the “late” onset trajectory group, but not the consistently “high” scorers, had higher global white matter diffusivity values than “low” symptom scorers, this effect was not significant for FA. The volume of white matter hyperintensities in the “late” onset trajectory group was significantly higher than in the consistently “high” scorers.

**Figure 3.**
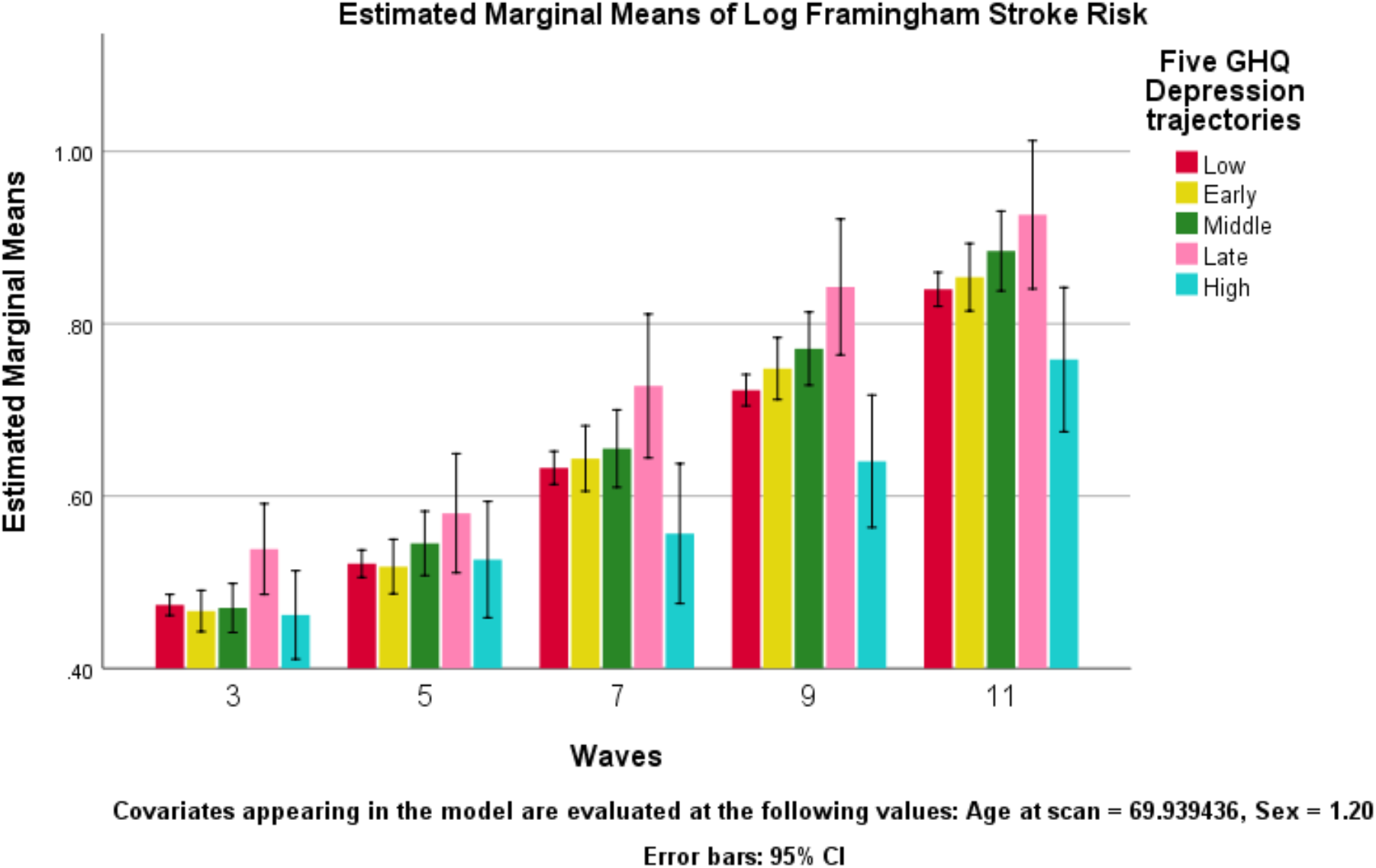
Framingham vascular (stroke) risks in estimated GHQ trajectory groups. The “late” symptom onset group had higher vascular risks than the “low” symptom group in waves 3, 9, and 11. In contrast, the consistently “high” symptom group had similar vascular risk as the “low” symptom group during all waves. The “late” symptom and the consistently “high” symptom group had significantly different vascular risks in wave 9 and on average across all waves (Table 3).

**Figure 4:**
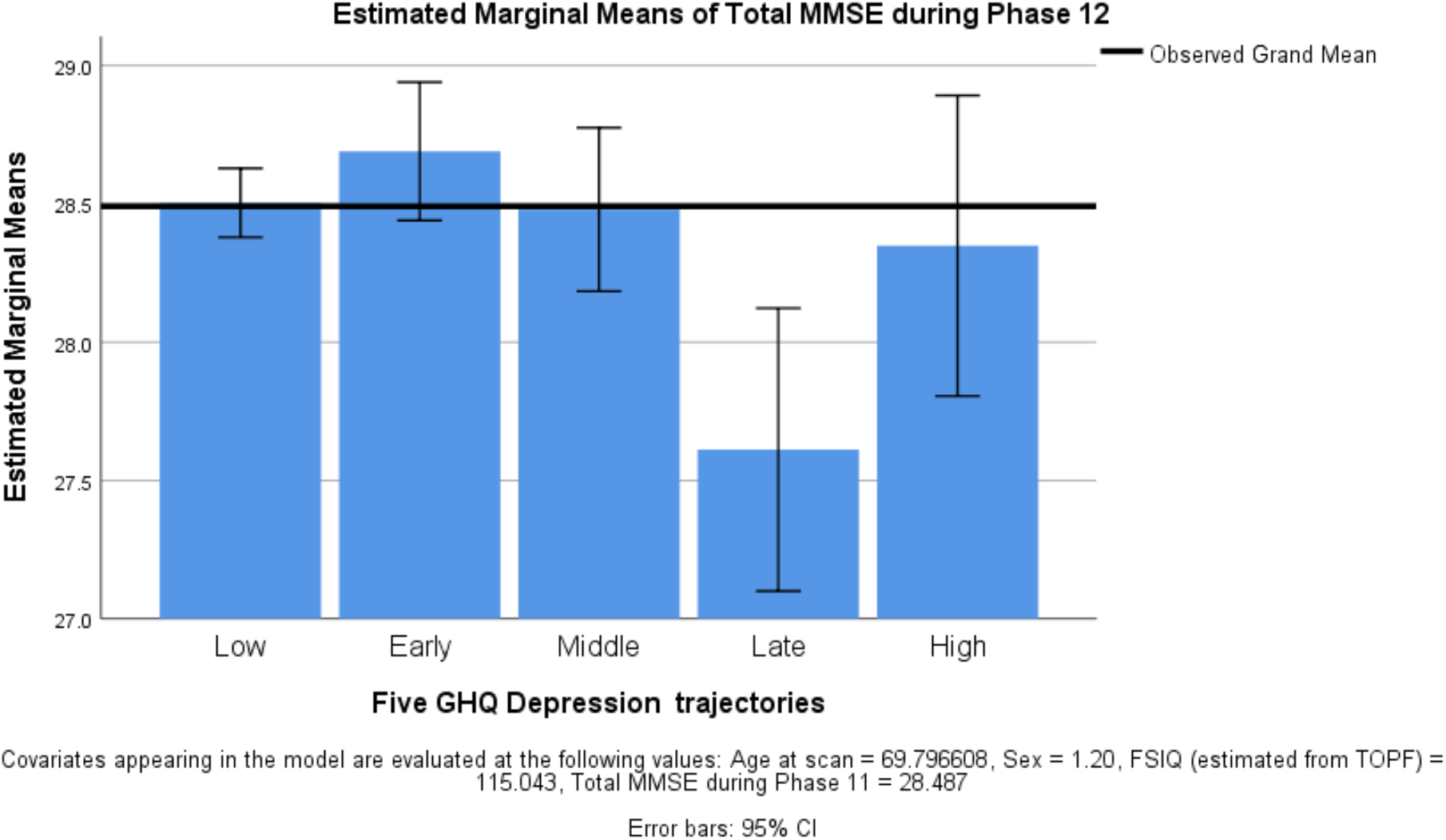
Comparison of MMSE score changes from Waves 11 to 12 between depression trajectory groups. After transformation of MMSE scores to Log (31-MMSE), the between-groups omnibus test was F_(4,714)_=3.704, p=0.005. After Bonferroni correction, the “Late” group had significantly lower MMSE scores at Wave 12 than the “Low” and “Early” symptom groups.

## Discussion

In this large group of community dwelling older adults, we identified five trajectories of depressive symptoms over a period of 28 years: those with low, early, middle, late and consistently high depressive symptoms. We hypothesized that a pattern depicting a late increase in depressive symptoms would be associated with decreased cognitive function and measures of adverse brain outcomes. In line with our hypothesis, participants in the late depressive symptoms group had a significantly poorer score on the TMT B-A, indicative of difficulties with executive function, increased white matter hyperintensities and increased vascular risk factors going back up to 22 years. We also found that participants in this trajectory group had reduced average MMSE scores over the 3.2 years between Waves 11 and 12, suggesting that a pattern of late-onset depressive symptoms might be associated with decreasing cognitive function.

The pattern of trajectories observed in our study, as well as the distribution of participants amongst them, is comparable with other studies of community-dwelling older adults (2). Similarly, our findings add to existing reports of cognitive decline, especially in executive function, as a characteristic of late-onset depression (7, 30). While some studies suggest that older adults may develop cognitive impairment as a result of experiencing depressive symptoms and the pathological correlates of depressive episodes or stress (31, 32), others argue that depressive symptoms are a prodromal feature of cognitive decline, reflecting a shared underlying pathology (3, 5, 7), or that depression may be a risk factor for cognitive impairment (33). The first hypothesis, not supported by our data, would predict the presence of poor cognitive performance in all four groups showing depressive symptoms. For the risk-factor hypothesis, also not supported by our data, one would expect the group with consistent high depressive symptoms to show the largest (‘dose dependent’) decline in cognitive function. Our findings, in line with others (2), support the hypothesis that late-onset depressive symptoms may be an early manifestation of cognitive decline. Furthermore, these findings highlight the value of screening patients presenting with depressive symptoms in late life for cognitive deficits.

Given the evidence pointing towards vascular disease as the link between depression and dementia (for review, see Byers and Yaffe (34)), and consistent with the greater vascular risk of the late onset group (Table 3, Figure 3), we had hypothesized that the propensity for cognitive decline observed in the late-onset trajectory group would be associated with shared brain pathologies and increased vascular risk factors. As predicted, the groups differed in white matter hyperintensity volume, a marker of microvascular changes (35), as well as mean diffusivity in white matter, suggesting impaired white matter structural integrity in the late onset group. Given that vascular factors are hypothesized to cause white matter damage (36), our findings are in line with the vascular depression hypothesis linking vascular risk factors to late-life depression (6). Contrary to our prediction, we did not find a significantly reduced fractional anisotropy in the group with late onset of depressive symptoms. Although we had a large sample of community-dwelling adults, the majority did not present with depressive symptoms, and only a small number of participants showed a consistently high or late-onset trajectory of depressive symptoms. Accordingly, the analysis may have been underpowered to estimate a relationship with these brain outcomes.

Kml is a non-parametric clustering method, which has proven useful in discerning clinical and behavioral trajectories in older adults (37, 38). Given its non-parametric nature, Kml does not test the fit between the partition found and a theoretical model. This is both a strength and a weakness. On one hand, this model-free approach allows us to derive more data-driven trajectories without the restriction of set parameters. On the other hand, without a model to be tested against, it is not possible to test the fit nor to calculate the likelihood of class assignments, as is done in model-based methods (e.g. latent class growth modelling). As with any trajectory analysis, it’s also important to recognize that assumptions are being made regarding the interval between measures of depressive symptoms, and we cannot discard the possibility that additional changes occurred between time-points. Other limitations in our study include the use of a shortened depressive symptom measure, the GHQ-depression sub-score, which may question the validity of its assessment of depressive symptoms. Nonetheless, previous studies have found that this measure is well suited for detecting depression in the general population (39, 40). In addition, the percentage of cases by Wave, as defined by the standard caseness criterion of the GHQ scale (Figure 2), was in line with the patterns observed in our trajectories. The proportions, if not the absolute values, for diagnostic frequencies (Table 1) determined by questionnaire and interview support this conclusion. Finally, since the Whitehall II cohort consists of British civil-servants recruited in the 1980s, a work-force that was then predominantly male, our sample was predominantly male. Given that there is a higher prevalence of depressive symptoms in females than males, arguably due to a combination of genetic, hormonal and psychosocial gender differences (41), our findings although adjusted for sex may not generalize to both sexes. We did not have the appropriate sex distribution to explore this in our cohort, so it would be of interest for further studies to examine the interactions between sex, depressive symptoms and brain structures in the context of risk for cognitive decline.

Strengths of our study include a long follow-up period of almost 3 decades and a well-characterized sample, enabling us to comprehensively examine a list of variables as potential predictors and confounders of depressive symptom trajectories. Also of note, the use of 9 repeated measures of depressive symptoms, a higher frequency than most comparable studies (1), enabled us to estimate more complex trajectory patterns. It is worth noting that at the time of scan, participants in the late-onset and consistently high symptom groups did not differ in terms of severity of depressive symptoms, as evidenced by their CES-D and GHQ-depression scores. This highlights how valuable information can be extracted from trajectory analyses, as these two groups, with different prognoses for later cognitive outcome, could not have been distinguished cross-sectionally without knowledge of their history.

In conclusion, our study of trajectories of depressive symptoms over almost 3 decades indicates that a late emerging pattern of depressive symptoms is associated with increased vascular risk over 20 years, with impaired brain white matter structure and current executive dysfunction, but not poor memory. It also predicted further deterioration in MMSE scores over 3.2 years. Altogether, our findings suggest that tracking depressive symptoms over time may be a useful tool to identify “pre-clinical” phenotypes of cognitive and brain disorders in the community with different etiologies and prognoses.

## Data Availability

The study follows Medical Research Council data sharing policies (https://mrc.ukri.org/research/policies-and-guidance-for-researchers/data-sharing/). In accordance with these guidelines, data from the Whitehall II Imaging Substudy is accessible via the Dementias Platform UK (https://portal.dementiasplatform.uk/), through formal request to the data sharing committee (https://www.ucl.ac.uk/iehc/research/epidemiology-public-health/research/whitehallII/data-sharing).

## Acknowledgements

The study was supported by the MRC grants “Predicting MRI abnormalities with longitudinal data of the Whitehall II Sub-study” (PI KPE), ClinicalTrials.gov Identifier: NCT03335696, and “Adult Determinants of Late Life Depression, Cognitive Decline and Physical Functioning - The Whitehall II Ageing Study” (PI: MK). This research was supported by the NIHR Oxford Health Biomedical Research Centre. The Wellcome Centre for Integrative Neuroimaging is supported by core funding from the Wellcome Trust (203139/Z/16/Z). MK is supported by the MRC (MR/S011676/1, MR/R024227), NIH (R01AG056477, RF1AG062553), and the Academy of Finland. ASM is supported by grants from the National Institute on Aging, NIH (R01AG056477, RF1AG062553). CES is now a full-time employee of the Alzheimer’s Association. Work on the Whitehall Imaging Sub-study was funded by the UK Medical Research Council (G1001354), and the HDH Wills 1965 Charitable Trust (1117747). While working on this research study, EZs and SS were funded by the EU Horizon 2020 grant (732592).

## Supplementary Information

**Appendix 1.**
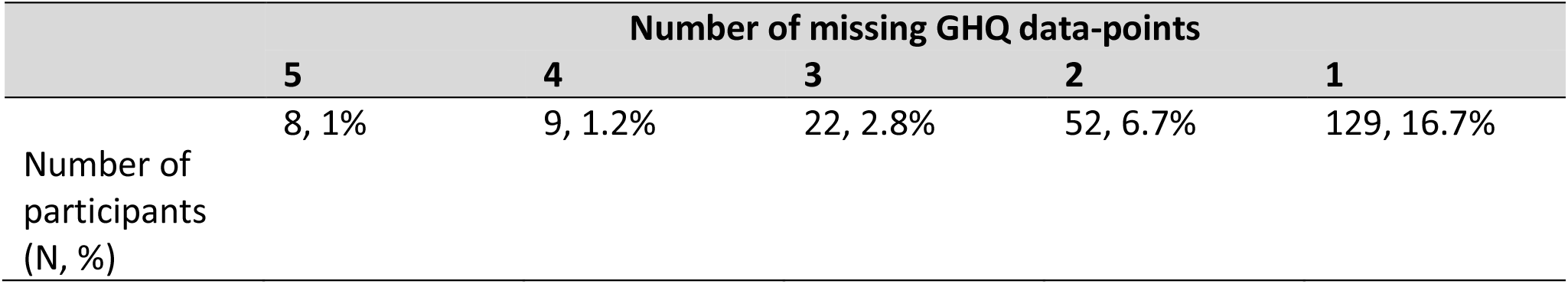
Overview of missing GHQ data.

|  | Number of missing GHQ data-points |  |  |  |  |
| --- | --- | --- | --- | --- | --- |
|  | 5 | 4 | 3 | 2 | 1 |
| Number of participants (N, %) | 8, 1% | 9, 1.2% | 22, 2.8% | 52, 6.7% | 129, 16.7% |

## References

1. Musliner KL, Munk-Olsen T, Eaton WW, Zandi PP (2016): Heterogeneity in longterm trajectories of depressive symptoms: Patterns, predictors and outcomes. Journal of affective disorders. 192: 199–211.

2. Mirza SS, Wolters FJ, Swanson SA, Koudstaal PJ, Hofman A, Tiemeier H, et al. (2016): 10-year trajectories of depressive symptoms and risk of dementia: a population-based study. Lancet Psychiatry. 3: 628–635.

3. Singh-Manoux A, Dugravot A, Fournier A, Abell J, Ebmeier K, Kivimaki M, et al. (2017): Trajectories of Depressive Symptoms Before Diagnosis of Dementia: A 28-Year Follow-up Study. JAMA Psychiatry. 74: 712–718.

4. Almeida OP, Hankey GJ, Yeap BB, Golledge J, Flicker L (2017): Depression as a modifiable factor to decrease the risk of dementia. Transl Psychiatry. 7:e1117.

5. Herrmann LL, Le Masurier M, Ebmeier KP (2008): White matter hyperintensities in late life depression: a systematic review. Journal of neurology, neurosurgery, and psychiatry. 79: 619–624.

6. Alexopoulos GS, Meyers BS, Young RC, Campbell S, Silbersweig D, Charlson M (1997): ‘Vascular depression’ hypothesis. Arch Gen Psychiatry. 54: 915–922.

7. Herrmann LL, Goodwin GM, Ebmeier KP (2007): The cognitive neuropsychology of depression in the elderly. Psychol Med. 37: 1693–1702.

8. Eraydin IE, Mueller C, Corbett A, Ballard C, Brooker H, Wesnes K, et al. (2019): Investigating the relationship between age of onset of depressive disorder and cognitive function. Int J Geriatr Psychiatry. 34: 38–46.

9. Castaneda AE, Suvisaari J, Marttunen M, Perala J, Saarni SI, Aalto-Setala T, et al. (2008): Cognitive functioning in a population-based sample of young adults with a history of non-psychotic unipolar depressive disorders without psychiatric comorbidity. Journal of affective disorders. 110: 36–45.

10. Brodaty H, Luscombe G, Peisah C, Anstey K, Andrews G (2001): A 25-year longitudinal, comparison study of the outcome of depression. Psychological medicine. 31: 1347–1359.

11. Marmot M, Brunner E (2005): Cohort profile: The Whitehall II study. Int J Epidemiol. 34: 251–256.

12. Filippini N, Zsoldos E, Haapakoski R, Sexton CE, Mahmood A, Allan CL, et al. (2014): Study protocol: the Whitehall II imaging sub-study. Bmc Psychiatry. 14.

13. Head J, Stansfeld SA, Ebmeier KP, Geddes JR, Allan CL, Lewis G, et al. (2013): Use of self-administered instruments to assess psychiatric disorders in older people: validity of the General Health Questionnaire, the Center for Epidemiologic Studies Depression Scale and the self-completion version of the revised Clinical Interview Schedule. Psychological medicine. 43: 2649–2656.

14. Reitan RM (1955): The relation of the trail making test to organic brain damage. J Consult Psychol. 19: 393–394.

15. Shapiro AM, Benedict RH, Schretlen D, Brandt J (1999): Construct and concurrent validity of the Hopkins Verbal Learning Test-revised. Clin Neuropsychol. 13: 348–358.

16. Folstein MF, Folstein SE, McHugh PR (1975): “Mini-mental state”. A practical method for grading the cognitive state of patients for the clinician. J Psychiatr Res. 12: 189-198.

17. Zhang Y, Brady M, Smith S (2001): Segmentation of brain MR images through a hidden Markov random field model and the expectation-maximization algorithm. IEEE Trans Med Imaging. 20: 45–57.

18. Smith SM (2002): Fast robust automated brain extraction. Human brain mapping. 17: 143–155.

19. Andersson J, Jenkinson M, Smith S (2007): Non-linear registration aka spatial normalisation. FMRIB Technical Report TR07JA2.

20. Griffanti L, Zamboni G, Khan A, Li L, Bonifacio G, Sundaresan V, et al. (2016): BIANCA (Brain Intensity AbNormality Classification Algorithm): A new tool for automated segmentation of white matter hyperintensities. Neuroimage. 141: 191–205.

21. Griffanti L, Jenkinson M, Suri S, Zsoldos E, Mahmood A, Filippini N, et al. (2018): Classification and characterization of periventricular and deep white matter hyperintensities on MRI: A study in older adults. Neuroimage. 170: 174–181.

22. Buysse DJ, Reynolds CF, 3rd, Monk TH, Berman SR, Kupfer DJ (1989): The Pittsburgh Sleep Quality Index: a new instrument for psychiatric practice and research. Psychiatry Res. 28: 193–213.

23. First MB, Gibbon M, Spitzer RL, Williams JBW (2002): Structured Clinical Interview for DSM-IV-TR Axis I Disorders (Research Version). In: Institute BRDNYSP, editor. New York.

24. Radloff LS (1977): The CES-D Scale: A Self-Report Depression Scale for Research in the General Population. Applied Psychological Measurement 1: 385–401.

25. Genolini C, Falissard B (2011): KmL: a package to cluster longitudinal data. Comput Methods Programs Biomed. 104:e112–121.

26. Genolini C, Alacoque, X., Sentenac, M., Arnaud C. (2015): kml and kml3d: R Packages to Cluster Longitudinal Data. Journal of Statistical Software. 65: 1–34.

27. RStudio Team (2016): RStudio: Integrated Development for R. Boston, MA.

28. Revelle W (2018): psych: Procedures for Personality and Psychological Research. 1.8.12 ed. Illinois, USA.

29. Wickham H, Sievert C (2016): ggplot2: elegant graphics for data analysis. Second edition. ed. Switzerland: Springer.

30. Taylor WD, Aizenstein HJ, Alexopoulos GS (2013): The vascular depression hypothesis: mechanisms linking vascular disease with depression. Mol Psychiatry. 18: 963-974.

31. Sapolsky RM, Krey LC, McEwen BS (1985): Prolonged glucocorticoid exposure reduces hippocampal neuron number: implications for aging. J Neurosci. 5: 1222–1227.

32. Marazziti D, Consoli G, Picchetti M, Carlini M, Faravelli L (2010): Cognitive impairment in major depression. Eur J Pharmacol. 626: 83–86.

33. Ownby RL, Crocco E, Acevedo A, John V, Loewenstein D (2006): Depression and risk for Alzheimer disease: systematic review, meta-analysis, and metaregression analysis. Arch Gen Psychiatry. 63: 530–538.

34. Byers AL, Yaffe K (2011): Depression and risk of developing dementia. Nat Rev Neurol. 7: 323–331.

35. Debette S, Markus HS (2010): The clinical importance of white matter hyperintensities on brain magnetic resonance imaging: systematic review and meta-analysis. Bmj. 341:c3666.

36. Zsoldos E, Filippini N, Mahmood A, Mackay CE, Singh-Manoux A, Kivimaki M, et al. (2018): Allostatic load as a predictor of grey matter volume and white matter integrity in old age: The Whitehall II MRI study. Sci Rep. 8: 6411.

37. Chamberlain AM, St Sauver JL, Jacobson DJ, Manemann SM, Fan C, Roger VL, et al. (2016): Social and behavioural factors associated with frailty trajectories in a population-based cohort of older adults. BMJ Open. 6:e011410.

38. Demmelmaier I, Dufour AB, Nordgren B, Opava CH (2016): Trajectories of Physical Activity Over Two Years in Persons With Rheumatoid Arthritis. Arthritis Care Res (Hoboken). 68: 1069–1077.

39. Koeter MW (1992): Validity of the GHQ and SCL anxiety and depression scales: a comparative study. Journal of affective disorders. 24: 271–279.

40. Lundin A, Hallgren M, Theobald H, Hellgren C, Torgen M (2016): Validity of the 12-item version of the General Health Questionnaire in detecting depression in the general population. Public Health. 136: 66–74.

41. Kuehner C (2017): Why is depression more common among women than among men? Lancet Psychiatry. 4: 146–158.

